# Tabular Foundation Model for Breast Cancer Prognosis using Gene Expression Data

**DOI:** 10.1101/2025.10.03.25337265

**Authors:** Tuyen Vu, Ha X. Tran, Xiaomei Li, Lin Liu, Jiuyong Li, Sindy Pinero, Jia Tina Du, Thuc D. Le

## Abstract

Accurate and robust survival prediction is essential for personalised breast cancer prognosis and treatment decision-making. However, existing machine learning–based survival models often lack stability under cohort heterogeneity and distribution shift, while tabular foundation models typically do not support censored time-to-event data. This study aims to develop a foundation-model-based survival framework that explicitly handles right censoring and enables stable deployment across heterogeneous cohorts without task-specific retraining.

We propose TabSurv, a foundation-model-based framework for survival analysis that adapts pretrained tabular foundation models to censored time-to-event prediction. Built on TabPFN, TabSurv operates via in-context learning without gradient-based training or hyper-parameter tuning. To address censoring, TabSurv introduces a two-stage, censoring-aware strategy. In the first stage, the model learns survival time relationships from uncensored observations and imputes survival outcomes for censored patients under a non-informative censoring assumption. In the second stage, imputed and observed outcomes are combined to reconstruct the learning context for final inference, reformulating survival analysis as a regression task. For treatment recommendation, TabSurv estimates individualised counterfactual survival risks using gene expression features restricted to biologically validated cancer driver genes.

TabSurv was evaluated on twelve breast cancer gene expression cohorts under in-distribution and out-of-distribution settings. It achieved competitive or superior prognostic performance compared with seven established survival models and demonstrated the highest predictive stability under cohort heterogeneity. In treatment recommendation experiments on METABRIC, patients whose treatments aligned with TabSurv’s recommendations exhibited significantly improved long-term survival.

TabSurv provides an efficient and stable foundation-model-based approach to censored survival analysis, supporting robust prognosis and personalised treatment recommendation in heterogeneous breast cancer cohorts.

## 1 Introduction

Breast cancer prognosis is essential for guiding clinical decision-making and tailoring treatment strategies to improve patient survival. In routine clinical practice, breast cancer prognosis has traditionally been assessed using ana-tomical staging systems, most notably the Tumour – Node – Metastasis (TNM) classification, which stratifies patients based on primary tumour size, lymph node involvement, and the presence of distant metastases. ^1,2^ While these clinical indicators enable effective population-level risk stratification, patients with similar tumour stage, grade, and nodal status can experience substantially different survival outcomes and treatment responses due to underlying breast cancer heterogeneity. ^3^ This diversity arises from distinct molecular processes governing tumour initiation, progression, and therapeutic resistance.

To better capture this heterogeneity, increasing attention has focused on the use of transcriptomic data for breast cancer prognosis at the molecular level. Gene expression profiles offer a more detailed characterisation of tumour biology and have been shown to improve risk stratification beyond conventional clinical factors. ^4,5^ In this context, survival analysis provides a principled statistical framework for modelling time-to-event outcomes, such as overall survival, disease-free survival, and recurrence risk, while appropriately accounting for censoring. ^6,7^ Censoring occurs when the true time-to-event is unknown beyond the observation period, a condition that traditional regression models are not equipped to handle. ^8,9^ The development of robust survival models capable of integrating high-dimensional molecular data represents both a critical challenge and a significant opportunity for improvingimproving personalised prognosis and therapeutic decision-making in breast cancer.

Numerous machine-learning-based survival models have been proposed for breast cancer prognosis, which can be broadly categorised into three groups: tree-based methods, such as Random Survival Forests (RSF); ^10^ discrete-time models, including Logistic Hazard , ^11^ Piecewise Constant Hazard (PCHazard) , ^11^ Probability Mass Function (PMF) , ^11^ Multi-Task Logistic Regression (MTLR) , ^12^ and DeepHit; ^13^ and neural network approaches, exemplified by DeepSurv. ^14^ These methods move beyond the classical Cox Proportional Hazards model ^6^ by capturing non-linear relationships in high-dimensional clinical and molecular data. However, most existing approaches implicitly assume that the training and test datasets are drawn from the same distribution, an assumption that is frequently violated in real-world clinical settings.

In practice, survival analysis models are often applied across cohorts that differ substantially in patient demographics, disease stage distributions, and gene expression measurement platforms. ^15,16^ Such distribution shifts can lead to a marked degradation in predictive performance and unreliable risk stratification, even for models that demonstrate high performance during internal validation. Ensuring stability, defined as the maintenance of consistent predictive accuracy across heterogeneous datasets, is therefore essential for the clinical translation of survival models. To address this, several strategies have been investigated, including multi-cohort training coupled with robust feature selection ^17,18^ and transfer learning frameworks designed to extract domain-invariant representations from independent pan-cancer datasets. ^19,20^ While effective in some settings, these approaches rely on the availability of multiple large, well-annotated survival datasets, which are costly and time-consuming to collect due to the requirement for long-term follow-up and comprehensive molecular profiling.

Recent advances in foundation models for tabular data enable pre-training on large collections of synthetic or real-world tabular tasks, often comprising millions of samples, which improves data efficiency, robustness, and stability under distribution shift. ^21,22^ Foundation models have demonstrated strong performance and generalisation across a wide range of predictive tasks. However, most existing foundation models are designed for fully observed outcomes and do not explicitly account for censored data, which is intrinsic to time-to-event modelling. ^8^ Although some foundation-model-based approaches for time-to-event prediction have recently been proposed, such as Many Outcome Time Oriented Representations (MOTOR), ^23^ there remains limited research on adapting general-purpose tabular foundation models to survival settings involving censoring. In contrast, classical survival methods explicitly model time-to-event outcomes and incorporate censoring during estimation, a capability that has not yet been systematically integrated into foundation-model-based frameworks.

These shortcomings identify three primary research gaps: (1) the need for survival models with higher predictive accuracy; (2) the requirement for robust prediction across heterogeneous breast cancer cohorts; and (3) the development of foundation-model-based methods that handle censoring while remaining easy to deploy without extensive training data or hyperparameter tuning.

To address these gaps, we propose TabSurv, a foundation-model-based framework for survival analysis that is specifically designed for tabular time-to-event data with censoring. TabSurv is built upon TabPFN (Tabular Prior-Data Fitted Network), ^22^ a transformer-based foundation model pretrained on millions of synthetic datasets. A key feature of TabPFN is in-context learning, which allows the model to instantly adapt to a new clinical cohort by using the provided examples as a reference, much like a clinician applying prior medical knowledge to a specific new case without needing to ”re-learn” medicine from scratch. Unlike traditional machine learning, this approach requires no gradient-based retraining or complex models hyperparameter tuning to adapt to new tasks. TabPFN achieves strong generalisation and data efficiency by approximating Bayesian posterior predictive inference, a method that formally updates the model’s prior knowledge with new evidence to make predictions. This evidence-based approach makes it particularly well-suited for survival prediction in settings with limited or heterogeneous clinical data.

TabSurv includes two main stages, censoring-aware learning strategy designed to adapt the foundation model to survival analysis. The first stage involves training TabSurv exclusively on uncensored observations. Under the assumption of non-informative censoring, where the censoring mechanism is independent of survival time conditional on the covariates, this approach provides a statistically valid foundation for estimating survival outcomes without introducing systematic bias. This initial model then estimates counterfactual survival times for censored patients: the durations expected had censoring not occurred, restricted by the requirement that they exceed the observed censoring point. In the second stage, these imputed times are merged with observed data to form a reconstructed training context, providing the basis for survival predictions on unseen patients. This two-stage framework allows TabSurv to incorporate the full dataset while remaining consistent with the established properties of the censoring mechanism. By formulating survival prediction as a regression problem, TabSurv provides a computationally efficient approach to breast cancer prognosis across diverse datasets, bypassing the need for iterative model retraining or extensive hyperparameter tuning.

Extending its utility to clinical decision support, TabSurv provides a framework for personalised treatment recommendation. Since its underlying foundation model (Tab-PFN) was pretrained on synthetic data derived from structural causal models, ^22^ its performance is maximised when input features reflect meaningful biological mechanisms rather than spurious correlations. We therefore restrict the genomic inputs to a curated set of biologically validated cancer driver genes, often referred to as causal genes, to reduce noise, improve interpretability, and enhance robustness. By estimating individualised counterfactual survival risks under alternative scenarios, TabSurv identifies the most favourable treatment pathway for each patient. Empirical evaluation confirms that patients whose actual treatment aligned with TabSurv’s recommendations exhibited significantly higher survival probabilities than those who received alternative therapies.

The main contributions of this paper are:

1. We developed TabSurv, a novel tabular foundation-model-based method, adapted for survival analysis through a two-stage censoring-handling mechanism.
2. The demonstration of TabSurv’s high efficacy in breast cancer prognosis using gene expression data. The model is validated across twelve diverse breast cancer datasets for both in-distribution and out-of-distribution scenarios, and benchmarked against seven state-of-the-art survival models, highlighting its robustness, efficiency and stability.
3. By integrating biologically validated driver genes into the feature selection process, TabSurv facilitates personalised treatment recommendations. Our experimental results demonstrate that patients whose treatments aligned with TabSurv’s recommendations have higher mean survival time compare to those whose did not follow recommendations, significantly outperforming seven baseline methods.

## 2 Methodology

### 2.1 Overview of the TabSurv Framework

Figure 1 illustrates the TabSurv framework, a foundation-model-based system for breast cancer prognosis and personalised treatment recommendation. TabSurv extends a pre-trained tabular foundation model, TabPFN, by introducing a two-stage, censoring-aware inference pipeline that accommodates right-censored data without requiring task-specific optimisation. In the first stage, the model uses uncensored samples as in-context references to map the relationship between covariates and survival outcomes, generating survival outcomes for censored individuals. These estimates are merged with observed events to form a reconstructed dataset with complete survival information. In the second stage, this reconstructed dataset provides the necessary context for final inference. This process allows for robust survival prediction by incorporating the full cohort distribution and the imputed time-to-event outcomes. The architecture integrates prognostic risk stratification with therapeutic decision support, providing both individualised risk assessments and estimations of counterfactual treatment outcomes within a single foundation-model framework.

**Figure 1:**
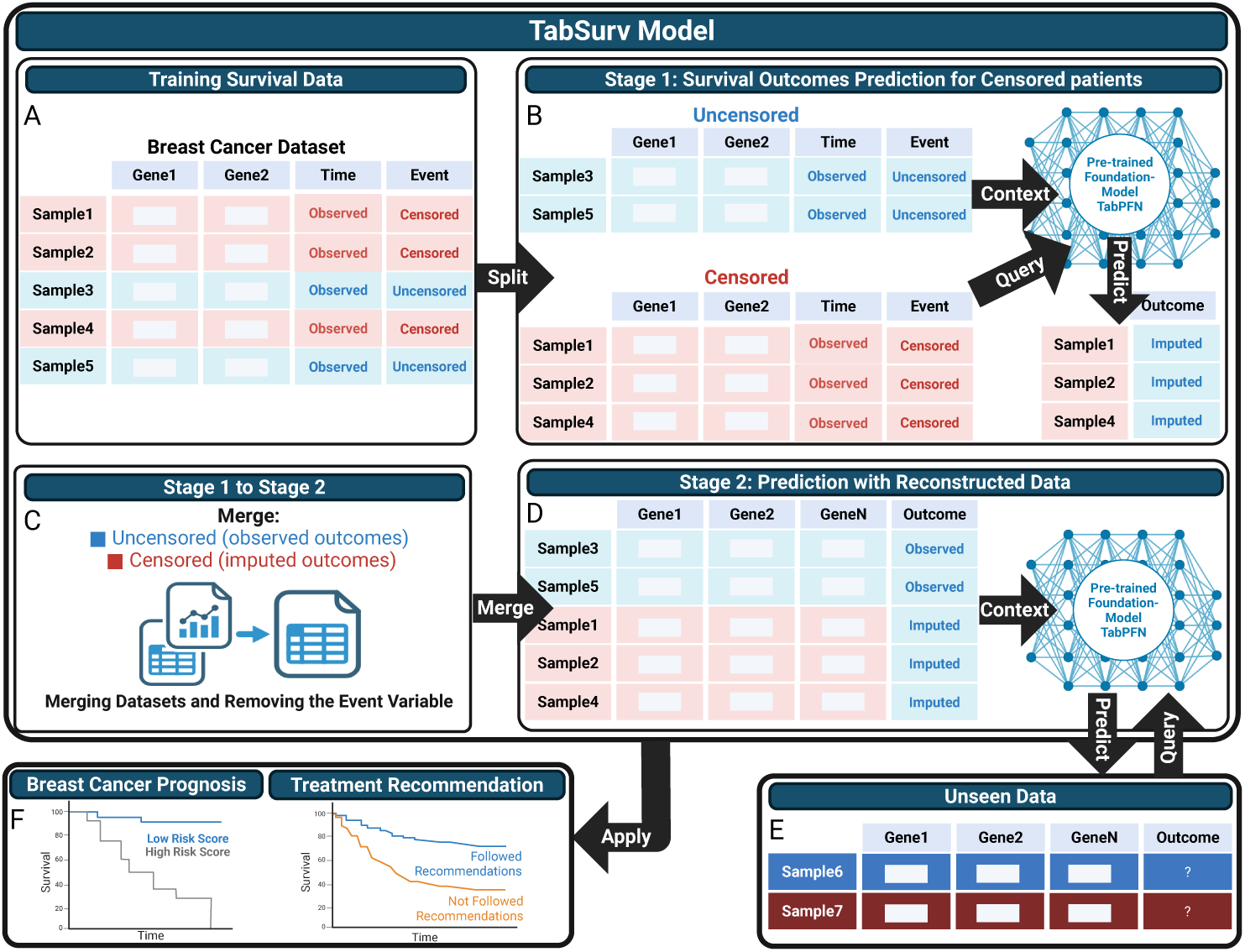
Overview of the TabSurv framework for breast cancer prognosis and treatment recommendation. (A) Input survival dataset containing genomic features with a mixture of uncensored (observed event times) and right-censored outcomes. (B) Stage 1 Survival outcomes prediction, where uncensored samples are used as in-context references for the pre-trained foundation-model-based, TabPFN, to learn covariate–survival relationships and to generate event-time predictions for censored individuals. (C) Dataset reconstruction by merging observed survival times from uncensored samples with imputed event times for censored samples. (D) Stage 2 prediction using the reconstructed dataset as context, enabling robust survival prediction from the full covariate distribution. (E) Application to unseen patients without task-specific retraining. (F) Downstream clinical applications, including breast cancer prognosis and personalised treatment recommendation evaluated using Kaplan–Meier survival curves for patients who followed or did not follow the model’s recommendations.

### 2.2 The Workflow of TabSurv

The TabSurv workflow employs a two-stage, censoring-aware procedure built upon a pre-trained foundation model (TabPFN) for both survival prediction and downstream treatment recommendation. The primary objective is to utilise in-context learning to handle right-censored data efficiently, bypassing the need for gradient-based retraining or hyperparameter tuning. ^22^ The complete architecture is illustrated in Figure 1 and is detailed through the following stages.

- **Stage 1: Survival Outcomes Estimation for the Censored Subset.** Given a breast cancer survival dataset with covariates *X*, observed follow-up times *T* , and an event indicator *δ* ∈ {0, 1}, the cohort is first partitioned into two subsets: uncensored patients (*δ* = 1), for whom true event times are observed, and censored patients (*δ* = 0), for whom only partial time information is available. The uncensored subset provides reliable supervision and serves as the initial reference context for model inference.

During the initial stage, the frozen TabPFN foundation model performs in-context regression by conditioning exclusively on uncensored samples, which serve as the context set. Instead of undergoing gradient-based training, the model maps the relationship between covariates and survival times directly from these provided examples. This model is then applied to the censored patients to estimate survival outcomes. To maintain consistency with the known follow-up data, these estimates are taken as the maximum of the predicted time and the observed censoring time. This ensures that no imputed survival time is shorter than the actual recorded follow-up. This step transforms censored observations into pseudo-complete samples while adhering strictly to the censoring mechanism.

- **Stage 2: Final Inference via Reconstructed Context.** The original uncensored samples are merged with the censored samples (now assigned imputed event times) to form a reconstructed dataset. This augmented cohort provides a comprehensive representation of the covariate distribution, capturing the characteristics of the entire population rather than just the uncensored subset.

In the subsequent stage, the frozen TabPFN is applied in an in-context manner, utilising the reconstructed dataset as the context set. By conditioning on both observed and imputed survival outcomes, the model benefits from broader coverage of the feature space. This architecture achieves more stable survival predictions for independent cohorts by utilising the collective data distribution of the entire study population to inform individual inferences.

- **Prognostic Assessment and Treatment Recommendation.** The TabSurv model generates individualised survival predictions that serve as a direct tool for breast cancer prognosis. Beyond risk stratification, the framework facilitates treatment recommendation by simulating counterfactual survival risks across alternative therapies. By duplicating patient profiles while holding all other covariates constant, the model estimates the likely outcome for each treatment arm. Under the assumption of no unmeasured confounding, where all variables influencing both treatment assignment and survival are captured in the model, these estimates allow for the identification of the most favourable therapy for each patient. The clinical utility of these recommendations is validated by comparing survival probabilities between patients whose actual treatment aligned with the model’s suggestion and those whose treatment did not, using Kaplan–Meier analysis.

### 2.3 Evaluation Metrics

To evaluate TabSurv for breast cancer prognosis and treatment recommendation, we adopt three complementary evaluation metrics addressing accuracy, robustness, and clinical utility. Prognostic performance is assessed using the Concordance Index (C-index), ^24^ which measures the agreement between predicted risk scores and observed time-to-event outcomes while accounting for censoring. The stability of TabSurv is evaluated by examining the consistency of prognostic performance across multiple in-distribution and out-of-distribution datasets, reflecting robustness under distribution shift. For treatment recommendation, a case is categorised as *Followed* if the therapy actually received by a patient in the observed dataset matches the model’s recommendation, and as *Not Followed* otherwise. This stratification allows for an evaluation of whether concordance with the model’s suggestions is associated with superior survival outcomes. Kaplan–Meier (KM) survival curves ^25^ and Restricted Mean Survival Time (RMST) ^26^ are employed to compare long-term survival between the *Followed* and *Not Followed* groups, providing a clinically grounded validation of the recommendation strategy’s potential utility.

#### 2.3.1 Concordance Index

The C-index ^24^ is a standard metric in survival analysis for evaluating the discriminative performance of prognostic models. It quantifies a model’s ability to correctly rank patients according to their risk of an event (e.g., disease recurrence or mortality). Formally, for a pair of patients (*i, j*), let *Y* denote the true survival time and *r* denote the predicted risk score. A pair is considered concordant if the model assigns a higher risk to the individual who experiences the event earlier; specifically, if *Y_j_ < Y_i_*, then *r_j_ > r_i_*.

In the presence of right-censored data, where the true survival time may be unobservable, we utilise the observed follow-up time *T_i_* and event indicator *δ_i_* (where *δ_i_* = 1 denotes an event and *δ_i_* = 0 denotes censoring). The C-index is calculated by considering all comparable pairs where the ordering of events is known with certainty: ^24,27]^

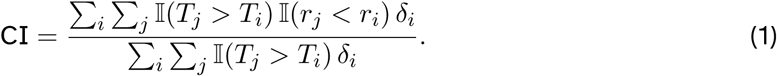

where I(·) is the indicator function. The index ranges from 0 to 1, where 0.5 represents chance and 1.0 indicates perfect concordance.

As TabSurv is designed to predict survival time *t̃* rather than a dimensionless risk score, we define the predicted risk as the additive inverse of the predicted survival time (*r_i_* = −*t̃i*). This transformation ensures compatibility with the standard C-index formulation, as a shorter predicted survival time correctly corresponds to a higher predicted risk. High C-index values in this context demonstrate the model’s capacity to accurately rank breast cancer patients according to their prognostic outlook.

#### 2.3.2 Stability Score

While predictive accuracy is essential, a clinically useful prognostic model must also be robust and generalisable to different datasets that may differ in sample size, feature distribution, and outcome prevalence. To capture this notion of robustness, we introduce stability metrics that quantify the consistency of model performance across multiple independent test datasets. Consider a collection of *M* test datasets, denoted as *D*^1^*, D*^2^*, …, D^M^* . Let *CI^m^* be the C-index obtained by the model on dataset *D^m^*. We define the stability of the model based on both the average performance and its variability across datasets as: ^28^

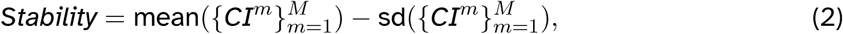

where mean(·) denotes the average C-index and sd(·) denotes the standard deviation across the *M* datasets. A higher stability score indicates that a model achieves not only strong average predictive performance but also low variability when evaluated on diverse datasets. Models with high stability are more reliable for clinical applications because they are less sensitive to dataset-specific factors such as cohort composition, data quality, or measurement variability.

This stability formulation is informed by the work of Kuang et al, 2018, ^28^ which evaluated model robustness by balancing predictive accuracy against error variability. By penalising excessive variance, this metric prioritises models that maintain consistent performance across heterogeneous clinical environments. In cancer prognosis, such stability is essential, as it indicates that a model may be reliably deployed across diverse patient populations without a substantial decline in predictive accuracy. Consequently, the combination of the C-index and the stability metric provides a rigorous assessment of both the accuracy and the generalisability of a prognostic method.

#### 2.3.3 Mean Survival Time

To evaluate the clinical utility of the treatment recommendation methods, we quantify survival differences between patient cohorts using the KM estimator ^25^ and a RMST based summary. ^26^ For each predictive model, patients are stratified into two groups according to concordance between the treatment they actually received and the model-recommended therapy: the *Followed* group, comprising patients whose administered treatment matched the recommendation, and the *Not Followed* group, consisting of patients whose treatment deviated from the model suggestion.

For each group, the KM estimator is used to obtain the non-parametric survival function *S*(*t*) = *P* (*T > t*), with survival curves plotted together with 95% confidence intervals to visualise longitudinal time-to-event dynamics. Statistical differences between survival distributions are assessed using the log-rank test.

To provide a clinically interpretable and robust quantitative comparison, we further summarise group-level survival using the RMST. Following the framework of Uno *et al.*, ^26^ RMST is defined as the expected survival time up to a pre-specified truncation time *τ* and is computed as the area under the KM survival curve,

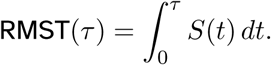

Unlike hazard-based metrics, RMST does not rely on the proportional hazards assumption and directly measures the average survival duration within a clinically time horizon. In this study, RMST provides an intuitive estimate of how long, on average, patients survive within the observation window when following versus not following model-recommended treatments. The difference in RMST between the *Followed* and *Not Followed* groups serves as a primary indicator of treatment recommendation effectiveness. A positive RMST difference indicates that adherence to the model’s recommendation is associated with a longer average survival time within the study follow-up time.

## 3 Experiment Results

### 3.1 Breast Cancer Prognosis with TabSurv

#### 3.1.1 Datasets

We used 12 breast cancer gene expression datasets from Li et al., 2025^17^ for this study. As shown in Table 1, these datasets were compiled from various public sources and include a total of 4,428 breast cancer patients. The *METABRIC* dataset is accessible publicly through the cBio-Portal for cancer genomics ^1^. *TCGA500* is a large-scale cancer genomics, provided by The Cancer Genome Atlas (TCGA) program^2^. Datasets including *MAINZ*, *TRANSBIG*, *UPP*, *UNT*, and *NKI* are obtained via Bioconductor^3^ using the data packages *breastCancerMAINZ*, *breastCancer-TRANSBIG*, *breastCancerUPP*, *breastCancerUNT*, and *breastCancerNKI*, respectively. The remaining datasets were retrieved from the Gene Expression Omnibus (GEO) database^4^.

**Table 1:** Summary statistics of the breast cancer datasets used in this study.

| Dataset | Sample size | Events | Event (%) | Min time | Max time |
| --- | --- | --- | --- | --- | --- |
| METABRIC | 1,980 | 647 | 32.68 | 0.00 | 29.60 |
| TCGA500 | 500 | 45 | 9.00 | 0.0027 | 17.91 |
| GEO | 736 | 349 | 47.40 | 0.00 | 18.52 |
| GSE6532 | 401 | 139 | 34.66 | 0.022 | 16.85 |
| GSE19783 | 100 | 43 | 43.00 | 0.69 | 10.62 |
| HEL | 115 | 25 | 21.74 | 0.00 | 5.00 |
| UNT | 133 | 28 | 21.05 | 0.17 | 14.53 |
| NKI | 320 | 109 | 34.17 | 0.02 | 18.35 |
| TRANSBIG | 198 | 62 | 31.31 | 0.34 | 29.60 |
| UK | 207 | 77 | 37.20 | 0.39 | 10.00 |
| MAINZ | 200 | 46 | 23.00 | 0.08 | 19.72 |
| UPP | 235 | 54 | 23.08 | 0.08 | 12.75 |

**Notes:** Sample size denotes the number of breast cancer patients in each dataset. Events correspond to the occurrence of the event of interest, including disease recurrence, metastasis, or death. Minimum and maximum times indicate the observed follow-up duration in years.

The clinical endpoints across these datasets vary: relapse-free survival (RFS) is used in *UPP*, *GSE6532*, *GEO*, *TCGA -500*, *METABRIC*, and *UK*; distant metastasis-free survival (DMFS) in *TRANSBIG*, *UNT*, *MAINZ*, and *NKI*; and death or DMFS in *HEL*. All survival times are recorded in years, with maximum durations ranging from 5 to 29.6 years. Events of interest include disease recurrence, metastasis, or death, with event rates ranging between 9.00% and 47.40%.

Due to differences in experimental batches, microarray/RNA -sequencing platforms, and preprocessing pipelines, the datasets exhibit heterogeneous distributions in both covariates and outcomes. This dataset shift complicates the development of models that generalise well to new, unseen cohorts. ^15,16^ To address this challenge, we propose a robust cancer prognosis model designed to perform reliably under distributional variability.

#### 3.1.2 Baselines

To evaluate the performance of TabSurv, we benchmark our framework against a diverse suite of established survival analysis models: DeepHitSingle (DeepHS), ^13^ DeepSurv, ^14^ Logistic Hazard (LH), ^29^ Multi-Task Logistic Regression (MTLR), ^12^ Piecewise Constant Hazard (PCHazard), ^29^ Probability Mass Function (PMF), ^11,29^ and Random Survival Forest (RSF). ^10^ These models are trained using both censored and uncensored clinical data to predict time-to-event outcomes. The benchmarking process includes standard preprocessing pipelines, such as one-hot encoding for categorical features, mean imputation for missing values, and the normalisation of continuous variables.

Performance is evaluated using the concordance index (C-index) as the primary measure of discriminative ability, together with a Stability Score defined in Section 2.3. For models that produce survival time estimates rather than risk scores, predicted times are transformed using the additive inverse (*r* = −*t*) to ensure compatibility with the C-index, where higher values indicate greater risk. This transformation preserves the required monotonic ordering and enables valid comparisons across different model architectures. To ensure a fair and comprehensive comparison across different modelling paradigms, we benchmarked the proposed framework TabSurv against a set of baseline survival models, including:

- **DeepHS**: ^13^ A fully parametric model that directly predicts the probability of an event occurring at each time point, optimised using a ranking loss and likelihood loss.
- **DeepSurv** : ^14^ A deep neural network that extends the Cox proportional hazards model to capture nonlinear relationships between features and survival risk.
- **LH**: ^29^ A discrete-time survival model based on logistic regression that models the conditional probability of failure at each time interval.
- **MTLR**: ^12^ A discrete-time survival model that treats survival prediction as a series of binary classification problems across time bins, jointly learned via logistic regression.
- **PCHazard**: ^29^ A piecewise constant hazard model implemented via neural networks, allowing for time-varying hazards across intervals.
- **PMF**: ^29^ A flexible discrete-time model that learns survival distributions directly by parameterising the probability mass function over time intervals.
- **RSF**: ^10^ A non-parametric ensemble method that constructs multiple survival trees using bootstrap samples and aggregates them to estimate cumulative hazard functions, representing the accumulated risk of an event occurring up to a specific time.

#### 3.1.3 Comparison of TabSurv with Baseline Methods Concordance Index

Table 2 presents the C-index performance of **TabSurv** compared with seven baseline survival models (*LogisticHazard*, *PMF*, *DeepHitSingle*, *PCHazard*, *MTLR*, *DeepSurv*, and *RSF*) in the out-of-distribution (OOD) scenario, where models are trained exclusively on the METABRIC dataset and evaluated on 11 independent breast cancer cohorts.

**Table 2:** Benchmarking survival models across 11 datasets under the out-of-distribution (OOD) <u>setting.</u>

| Dataset | TabSurv | DeepHS | DeepSurv | LH | MTLR | PCHazard | PMF | RSF |
| --- | --- | --- | --- | --- | --- | --- | --- | --- |
| TCGA500 | 0.6114 | 0.5187 | 0.5931 | 0.5602 | 0.5350 | **0.6450** | 0.5802 | <u>0.6178</u> |
| GEO | **0.6058** | 0.5033 | <u>0.5751</u> | 0.5508 | 0.5594 | 0.5775 | 0.5589 | 0.5732 |
| GSE6532 | **0.6554** | 0.6110 | <u>0.6375</u> | 0.5663 | 0.6519 | 0.6179 | 0.6091 | 0.6085 |
| GSE19783 | 0.5760 | 0.5170 | 0.5619 | **0.5907** | <u>0.5864</u> | 0.5589 | 0.5350 | 0.5678 |
| HEL | **0.6716** | 0.5100 | 0.4814 | 0.4588 | 0.4955 | 0.4647 | 0.4751 | <u>0.6685</u> |
| unt | 0.6736 | 0.5659 | **0.6768** | 0.6059 | <u>0.6718</u> | 0.6754 | 0.5984 | 0.6178 |
| nki | **0.6921** | 0.5954 | <u>0.6771</u> | 0.5675 | 0.6461 | 0.5994 | 0.6378 | 0.6705 |
| transbig | 0.6159 | 0.5828 | **0.6406** | 0.5878 | <u>0.6347</u> | 0.6000 | 0.5751 | 0.6206 |
| UK | **0.7087** | 0.6084 | 0.6673 | 0.5824 | <u>0.6850</u> | 0.6589 | 0.6789 | 0.6326 |
| mainz | **0.6663** | 0.5068 | <u>0.6230</u> | 0.5842 | 0.6009 | 0.6009 | 0.6133 | 0.6617 |
| upp | 0.6612 | 0.6689 | 0.6754 | 0.6034 | **0.6899** | 0.6436 | <u>0.6833</u> | 0.6837 |

**Notes:** LH, Logistic Hazard; PMF, Probability Mass Function; DeepHS, DeepHitSingle; MTLR, Multi-Task Logistic Regression; RSF, Random Survival Forest. Best-performing methods are shown in bold; second-best methods are underlined.

Across these datasets, TabSurv achieves the highest C-index on 6 out of 11 datasets, achieving the highest average performance and outperforming all baseline methods, suggesting robustness to distribution shift that typically degrade the performance of survival models.

Among competing models, Random Survival Forest emerges as the strongest baseline, obtaining the second-best score on six datasets. DeepSurv achieves the highest C-index on two datasets. While baseline models such as LogisticHazard, MTLR, and PCHazard perform reasonably on some datasets, their results vary considerably, reflecting their sensitivity to distribution shift.

Overall, a reliable survival model should yield a C-index above 0.5, indicating predictive concordance with observed event times. While most models surpass this threshold on the majority of datasets, TabSurv consistently delivers high concordance values and is the only method that maintains competitive performance across all OOD datasets without substantial degradation, underscoring the advantage of foundation-model inference and the proposed two-stage handling of censored data.

The experimental results of all models under the in-distribution (InD) setting across 12 datasets (Supplementary Table S1) show that TabSurv maintains more consistent performance across the majority of datasets.

#### Stability Metric

Table 3 presents the mean C-index, standard deviation, and stability score of each model across all datasets. The stability metric is defined as the mean C-index minus its standard deviation, capturing both predictive accuracy and robustness.

**Table 3:** Comparison of model stability across all evaluated survival models.

| Model | Mean C-index | Std. C-index | Stability |
| --- | --- | --- | --- |
| TabSurv | 0.6489 | 0.0409 | **0.6080** |
| DeepHS | 0.5626 | 0.0554 | 0.5072 |
| DeepSurv | 0.6190 | 0.0615 | 0.5575 |
| LH | 0.5689 | 0.0404 | 0.5285 |
| MTLR | 0.6142 | 0.0642 | 0.5500 |
| PCHazard | 0.6038 | 0.0581 | 0.5457 |
| PMF | 0.5950 | 0.0609 | 0.5341 |
| RSF | 0.6293 | 0.0387 | <u>0.5907</u> |

**Notes:** Mean C-index indicates average predictive discrimination across datasets. Std. C-index quantifies performance variability. The stability score is defined as *mean C-index minus standard deviation*, reflecting both accuracy and robustness. Higher values indicate more stable performance across heterogeneous cohorts.

TabSurv achieves the highest stability score (*Stability* = 0.608), outperforming all baseline methods. This demonstrates that TabSurv provides both accuracy and consistency across diverse datasets. Although some models, such as DeepHitSingle, show relatively low variability, their mean C-index values are lower, limiting their overall utility. These results suggest TabSurv achieves competitive accuracy with low variability.

### 3.2 Treatment recommendations for breast cancer patients with TabSurv

Precision oncology aims to personalise treatment selection, as therapeutic benefit varies markedly across patients due to differences in tumour biology, genomic heterogeneity, and treatment responsiveness. In this context, TabSurv provides a powerful engine for time-to-events prediction that can be used as a plug-in module for individualised treatment recommendation. Through the direct estimation of counterfactual risk scores and the integration of a censoring-aware training strategy, TabSurv facilitates the assessment of alternative treatment scenarios for individual patients.

#### 3.2.1 Counterfactual Risk Estimation for Treatment Recommendation

In addition to predicting survival time, TabSurv can be extended to estimate individualised risk scores in a manner comparable to classical survival models such as Random Survival Forests. This extension allows TabSurv to handle censoring while providing a unified risk-based framework for treatment recommendation.

Given a patient with feature vector *X*, TabSurv generates counterfactual predictions by modifying only the treatment variable while keeping all other clinical and genomic features fixed. Let *X*_(*t*)_ denote the same patient under treatment option *t* (e.g., chemotherapy or radiotherapy). The corresponding counterfactual risk score is given by;

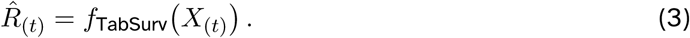

where *f*_TabSurv_ denotes the TabSurv risk-score prediction model, trained using a censoring-aware strategy.

In the treatment recommendation setting, competing treatment options are evaluated by comparing their predicted risk scores. For example, for chemotherapy, TabSurv estimates *R*^^^_Chemo=1_ and *R*^^^_Chemo=0_, and chemotherapy is recommended if *R*^^^_Chemo=1_*< R*^^^_Chemo=0_, indicating a lower predicted risk under treatment.

Similarly, for patients eligible for multiple treatment modalities, such as chemotherapy and radiotherapy, TabSurv compares *R*^^^_Chemo_ and *R*^^^_Radio_ and recommends the treatment associated with the lower predicted risk. This formulation naturally supports multi-arm treatment decision-making and provides a unified framework for counterfactual risk estimation.

The clinical effectiveness of the resulting treatment recommendations is evaluated by stratifying patients according to whether they followed the TabSurv recommendation, and comparing outcomes using KM survival curves and log-rank tests. This evaluation directly assesses whether lower predicted risk scores translate into improved observed survival.

#### 3.2.2 Selection of Biologically Important Genes

Feature selection constitutes an integral component of the proposed TabSurv framework. In addition to its two-stage censoring-aware training strategy, TabSurv explicitly incorporates biologically-informed feature selection to enhance robustness, generalisation, and interpretability when modelling high-dimensional genomic data.

This approach assumes that predictive performance is maximised when input features represent established causal mechanisms, specifically driver genes, rather than spurious correlations. To implement this, the framework restricts genomic inputs to a curated set of biologically validated cancer driver genes instead of utilising the full transcriptome. This constraint imposes a biological prior: prognostic signals that generalise across cohorts are likely concentrated within core oncogenic programmes. In contrast, broad transcriptomic measurements often contain redundant or platform-specific variation that may lead to overfitting and reduced out-of-distribution performance.

Specifically, we evaluated three established gene panels:

1. **Pereira-40 genes set**, derived from the comprehensive integrative genomic analysis of breast cancer by Pereira et al., ^30^ which identified recurrently mutated driver genes across a large breast cancer cohort.
2. **Mills-45 genes set**, based on clinically actionable and biologically relevant genes reported by Mills et al., ^31^ focusing on genes associated with targeted therapy response and breast cancer progression.
3. **Nik-94 genes set**, curated from the whole-genome sequencing study of 560 breast tumours by Nik-Zainal et al. ^32^ This panel comprises protein-coding genes with strong multistudy evidence of driver activity and established roles in breast tumorigenesis.

For each gene panel, TabSurv was trained and evaluated under identical experimental settings to ensure a fair comparison. The quantitative results, provided in Supplimentary Table S2, show that the *Nik-94 genes set* achieved the best performance in terms of the evaluation metrics for treatment recommendation, including survival differences between the Followed and Not Followed groups. Consequently, the *Nik-94 genes set* set was adopted as the default genomic input (DRIVER_CORE) for the main TabSurv treatment recommendation model.

This DRIVER_CORE set comprises 94 protein-coding genes with strong, multi-study evidence of driver activity. These genes have well-characterised roles in breast cancer biology: activating *PIK3CA* mutations occur in approximately 30–40% of cases and contribute to endocrine therapy resistance in HR+ breast cancer; ^33–36^ *FOXA1* encodes a pioneer transcription factor essential for the luminal phenotype, with alterations linked to endocrine resistance and disease progression; ^37,38^ cell-cycle regulators *CDKN1B* and *CCND1* are established determinants of proliferation and form the biological rationale for CDK4/6 inhibitor therapies; ^39^ the NOTCH pathway (*NOTCH1*, *NOTCH2*) regulates cancer stem cell maintenance and has been implicated in treatment resistance, particularly in triple-negative disease; ^40,41^ *CTNNB1* (*β*-catenin) mediates WNT signalling associated with epithelial-mesenchymal transition and metastatic potential; ^42^ and *NTRK3* represents an emerging therapeutic target. ^43^

Table 4 maps representative driver genes to higher-order biological programmes. By explicitly incorporating this biologically informed feature selection into the TabSurv pipeline, the framework reduces dimensionality, mitigates overfitting, and aligns the input space with the causal priors established during TabPFN’s pre-training, enabling more robust and reliable counterfactual risk estimation.

**Table 4:** Mapping from representative DRIVER_CORE genes to canonical breast cancer path- <u>ways and clinical implications.</u>

| Gene | Pathway | Clinical Interpretation |
| --- | --- | --- |
| *PIK3R1* | PI3K/AKT signalling | Survival and growth signalling; therapy response heterogeneity |
| *FOXA1* | Luminal lineage regulation | Differentiation and endocrine axis; subtype-specific prognosis |
| *CDKN1B* | Cell-cycle control | Proliferative constraint; recurrence risk and systemic therapy sensitivity |
| *CTNNB1* | WNT/ adhesion | Invasion and metastasis programmes; long-term outcome trajectories |
| *NOTCH1/2* | Notch signalling | Differentiation and stem-like programmes; resistance patterns |
| *NTRK3* | Receptor signalling | Targetable signalling axes; progression and differential therapy benefit |

#### 3.2.3 Evaluation Settings for Treatment Recommendation Experiments

We use the METABRIC dataset from the cBioPortal for Cancer Genomics, which includes 1,980 patients with clinical records (specifically chemotherapy and radiotherapy history), survival outcomes, and gene expression profiles. After excluding patients who received neither therapy or were treated with both, we retain a cohort of 1,063 individuals for our treatment recommendation analysis. The maximum observed survival follow-up time in this cohort is 20 years. After mapping to the DRIVER_CORE gene set, 72 genes remained available for the experimental setting.

Using the extended TabSurv framework, the model is trained on 60% of the 1,063 preprocessed samples from the METABRIC cohort and evaluated on the remaining 40%, with a fixed random seed of 42 to ensure reproducibility. For each patient in the test set, TabSurv estimates counterfactual risk scores for both treatment alternatives and recommends the intervention associated with the lower predicted risk.

The efficacy of the treatment recommendations is evaluated by stratifying patients into two cohorts: the *Followed* group (those whose received treatment matches the model’s recommendation) and the *Not Followed* group (those whose treatment deviated from the recommendation). KM survival curves are subsequently constructed to compare survival outcomes between these two cohorts, extending from baseline to the maximum observed follow-up period of 20 years.

Using the same dataset and setting for seven survival analysis models: DeepHitSingle , ^13^ DeepSurv , ^14^ Logistic Hazard , ^29^ Multi-Task Logistic Regression (MTLR) , ^12^ Piecewise Constant Hazard (PCHazard) , ^29^ Probability Mass Function (PMF) , ^11,29^ and Random Survival Forest (RSF) _. 10_

#### 3.2.4 Comparison of TabSurv Recommendation with Baseline Methods Using All Genes

Table 5 summarises the RMST, measured in years, for the *Followed* and *Not Followed* groups under the TabSurv recommendation and all baseline methods using the full gene set. The reported mean survival values correspond to the area under the Kaplan–Meier survival curve up to the pre-specified truncation horizon (Supplimentary Figure S2)., providing an interpretable estimate of average survival duration within the observed follow-up period. The difference in RMST (Δ Mean survival) quantifies the survival gain associated with adherence to each model’s recommended treatment.

**Table 5:** Comparison of mean survival time (years) between *Followed* and *Not Followed* groups across TabSurv R<u>ecommendation with Baseline Methods Using All Genes.</u>

| Model | Mean survival<br>(Followed<br>Group) | Mean survival<br>(Not Followed<br>Group) | $\Delta$ Mean<br>survival |
| --- | --- | --- | --- |
| TabSurv | 16.32 | 12.45 | **3.86** |
| DeepHS | 12.77 | 14.17 | -1.40 |
| DeepSurv | 14.20 | 12.69 | 1.51 |
| LH | 14.20 | 12.69 | 1.51 |
| MTLR | 13.58 | 13.71 | -0.12 |
| PCHazard | 14.27 | 12.33 | 1.93 |
| PMF | 13.89 | 13.53 | 0.36 |
| RSF | 12.69 | 14.20 | -1.51 |

$\Delta$ Mean survival denotes the difference between *Followed* Recommendation and *Not Followed* Recommendation groups.

Across all methods, TabSurv achieves the largest positive RMST difference, with patients in the *Followed* group exhibiting an average survival advantage of 3.86 years compared with those in the *Not Followed* group. This substantial margin indicates that treatments recommended by TabSurv are strongly aligned with improved long-term patient outcomes. In contrast, several baseline methods demonstrate markedly smaller RMST gains, while others exhibit negative differences, suggesting that following their recommendations is not associated with improved survival and may even be detrimental. For example, DeepHitSingle and RSF yield negative RMST differences, indicating longer average survival among patients whose treatments did not follow the model recommendations.

Overall, these findings highlight the superior clinical relevance of TabSurv’s treatment recommendations when evaluated using an RMST-based metric. Unlike hazard-based comparisons, RMST directly reflects expected survival time within a clinically meaningful horizon, reinforcing the conclusion that TabSurv provides more effective and actionable treatment guidance than existing baseline approaches.

#### 3.2.5 Comparison of TabSurv Recommendation with Baseline Methods Using Selected Genes

Table 6 reports the restricted mean survival time (RMST), expressed in years, for the *Followed* and *Not Followed* groups under the TabSurv recommendation framework and all baseline methods using biologically validated cancer driver genes. The difference in RMST (Δ Mean survival) corresponds to the area under the Kaplan–Meier survival curve (Supplimentary Figure S3) within the observed follow-up period.

**Table 6:**
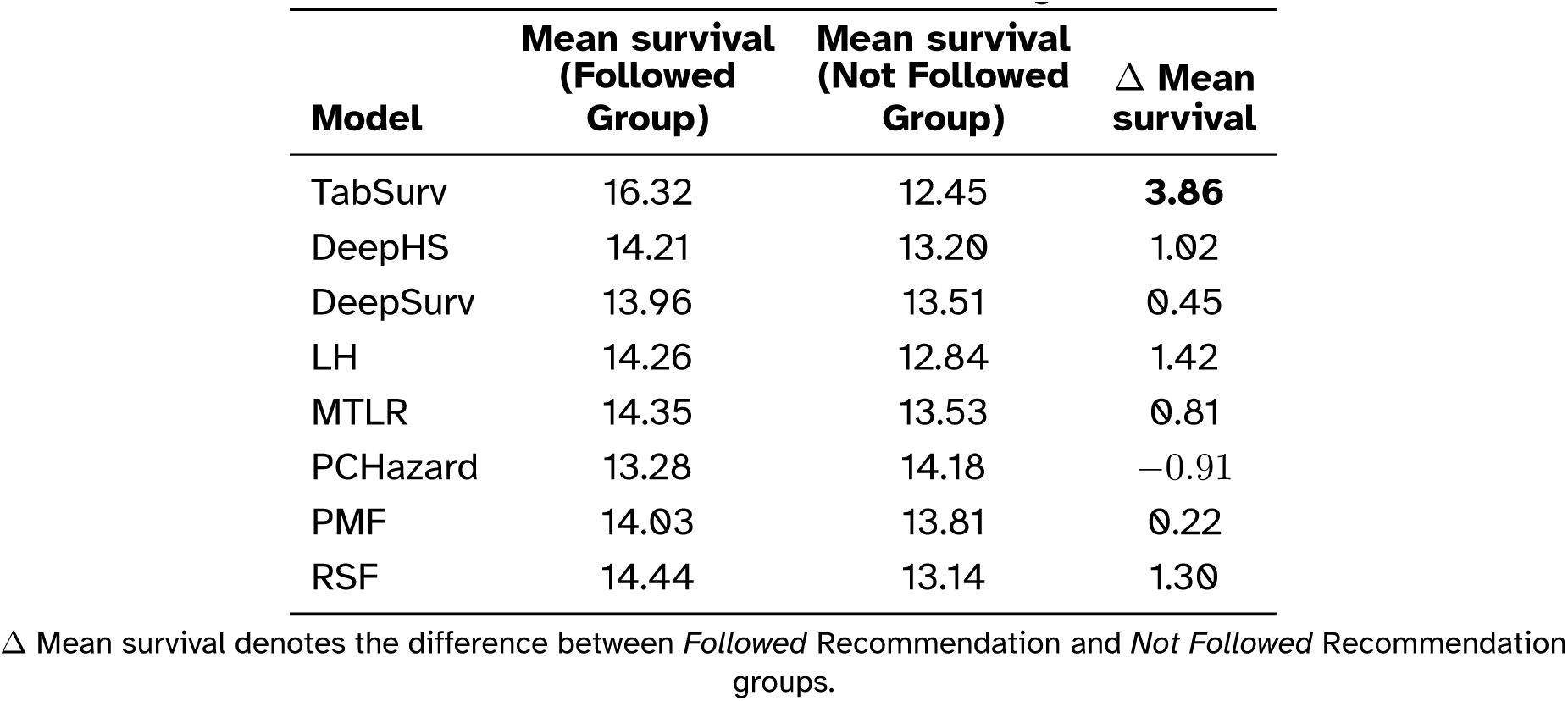
Comparison of mean survival time (years) between *Followed* and *Not Followed* groups across TabSurv R<u>ecommendation with Baseline Methods Using Selected Ge</u>nes.

The experimental results showed that the baseline methods did not outperform TabSurv. Overall, the performance of some baseline methods (DeepHS, MTLR, and RSF) under the reduced feature setting was better than when using all genes. However, although these models demonstrated some improvement, indicating slightly better alignment between their recommendations and long-term survival outcomes (positive Δ mean survival), these gains remained substantially smaller and less consistent than those observed for TabSurv.

These results indicate that TabSurv achieves higher treatment recommendation performance than the baseline methods, whether the baselines use the full gene set or a restricted subset of genes.

## 4 Discussion and Conclusion

TabSurv reformulates time-to-event prediction as a tabular regression task and employs incontext learning for survival inference, eliminating the need for gradient-based training or hyperparameter tuning. The two-stage censoring-handling workflow first learns survival-time outcomes from uncensored patients, then imputes pseudo-event times for censored cases, thereby constructing a reconstructed training context that improves predictive stability under incomplete follow-up. Evaluations across twelve breast cancer cohorts show that TabSurv achieves competitive or superior prognostic performance compared to RSF, DeepSurv, Logistic Hazard, PMF, DeepHit, PCHazard, and MTLR, while exhibiting the highest stability in out-of-distribution settings.

The stability metric, which summarises both mean concordance and dataset-to-dataset variability, reflects how consistently a method captures conserved tumour determinants across heterogeneous cohorts. The superior stability of TabSurv (Table 3) supports the hypothesis that driver-gene restriction aligns the model with conserved oncogenic pathways rather than dataset-specific signatures, providing resilience to variations in subtype mixture, treatment protocols, and endpoint definitions.

The framework extends survival prediction to treatment recommendation by generating counterfactual risk scores under alternative treatment assignments: given the same tumour state and patient context, which modality predicts a lower hazard trajectory? However, in observational cohorts, treatment assignment reflects clinical indication; consequently, counterfactual comparisons can be subject to residual confounding. The recommendations should therefore be interpreted as hypothesis-generating signals requiring validation through sensitivity analyses or independent clinical evidence. Our retrospective METABRIC analysis indicates that the framework recommendations align with superior survival outcomes. By restricting inputs to DRIVER_CORE genes, counterfactual estimates are grounded in biologically validated mechanisms, thus mitigating reliance on spurious correlations. The survival separation between patients who followed and those who did not follow the recommendations provides indirect evidence of clinical utility, suggesting that the recommendations capture latent differences in tumour behaviour and treatment response. TabSurv was the only method to demonstrate consistent benefit across all time horizons, whereas the baseline methods showed inconsistent performance patterns that may reflect overfitting to specific survival windows.

The TabSurv framework establishes a foundation that can be extended to additional data modalities and clinical settings. Multi-omics integration combining expression with mutation profiles, copy number alterations, and methylation data would capture complementary tumour biology, ^44,45^ while spatial transcriptomics or single-cell profiling would incorporate microenvironment effects. ^46,47^ The modular architecture of TabSurv easily supports these extensions, as well as competing risk modelling, calibrated survival probabilities with uncertainty quantification, and additional treatment modalities. Based on the positive retrospective results reported here, prospective validation in a randomised trial comparing TabSurv-guided selection with standard care represents a natural next step toward clinical translation.

Pathway enrichment analysis confirms that the DRIVER _CORE gene set captures biologically and clinically relevant tumour programmes. Direct enrichment for the KEGG breast cancer pathway confirms the disease-specific relevance, while enrichment for endocrine resistance aligns with the clinical importance of hormone therapy response in breast cancer prognosis. GO enrichment for the G1/S phase transition includes key cell-cycle genes such as *CDKN1B* and *CCND1*, which establish determinants of proliferative capacity and prognosis. Enrichment for gland development is consistent with mammary-specific biology and lineage regulators such as *FOXA1*. Significant enrichment of *PI3K* signalling in both GO and KEGG analyses supports the mechanistic relevance of *PIK3CA* and *PIK3R1* discussed in Section 3.2.2. The enrichment for radiation response has direct clinical relevance, given that radiotherapy is one of the treatment modalities in the TabSurv recommendation framework, suggesting that the DRIVER_CORE genes capture the variation relevant to distinguishing patients who may benefit more from radiotherapy than chemotherapy. The cross-cancer enrichment observed in KEGG (melanoma, prostate, hepatocellular, gastric) reflects conserved oncogenic programmes shared across malignancies; rather than constituting a limitation, this may contribute to TabSurv’s robustness by capturing fundamental tumour biology rather than breast-cancer-specific artefacts. The complete enrichment results are available in SM-GO and SM-KEGG.

The DRIVER_CORE gene set represents a coherent collection of core cancer mechanisms that control proliferation, survival signalling, lineage regulation, and therapy resistance, providing a mechanistic basis for TabSurv’s predictions. By integrating an accurate prognosis with treatment selection within a single foundation-model framework, TabSurv demonstrates that a biologically grounded, training-free survival analysis can match or surpass conventional approaches. The extensibility of the framework to multi-omics data, additional treatment modalities, and prospective clinical settings establishes TabSurv as a step toward translating foundation models into precision oncology practice.

## 5 Code availability

Codes and public datasets are available for public access on GitHub at https://github.com/vntuyen/tabsurv TabSurv was implemented in Python using the TabPFN library (version 6.3.0)^5^.

Baseline survival models were implemented in Python 3.13. Specifically, Logistic Hazard, Probability Mass Function, DeepHitSingle, PCHazard, MTLR, and DeepSurv were implemented using the pycox library (version 0.3.0)^6^. ^29^ Random Survival Forests (RSF) were implemented using the scikit-survival library (version 0.25.0)^7^. ^48^ Detailed configuration parameters and hyperparameter settings for each model are provided in the Github repository.

## Data Availability

All data produced in the present study are available upon reasonable request to the authors

https://www.cancer.gov/tcga

https://www.ebi.ac.uk/ega/

## Footnotes

1 https://www.cbioportal.org/study/summary?id=brca_metabric

2 https://www.cancer.gov/tcga

3 https://bioconductor.org/

4 https://www.ncbi.nlm.nih.gov/geo/

5 https://github.com/PriorLabs/TabPFN

6 https://github.com/havakv/pycox

7 https://scikit-survival.readthedocs.io/

