## Supplementary Material for "Tabular Foundation Model for Breast Cancer Prognosis using Gene Expression Data"

#### 1 Pathway Enrichment Analysis

To assess the biological coherence of the feature set used by TabSurv, we performed a pathway enrichment analysis on the 94 DRIVER_CORE genes. Of these, 90 genes were successfully mapped to Entrez gene identifiers using the org.Hs.eg.db annotation database [3] and used for subsequent analysis. The enrichment of Gene Ontology (GO) [1] was carried out in terms of the biological process, and the enrichment of the KEGG [4] pathway was carried out, with adjusted p-values using the Benjamini-Hochberg method [2] (adjusted $p < 0.05$ considered significant).

##### 1.1 Gene Ontology Enrichment

GO Biological Process analysis identified 1,453 significantly enriched terms (adjusted $p < 0.05$). The most enriched processes were gland development (26 genes; adjusted $p = 6.02 \times 10^{-18}$), regulation of the cell cycle phase transition (26 genes; adjusted $p = 7.39 \times 10^{-18}$) and the G1/S phase transition of the cell cycle (22 genes; adjusted $p = 8.46 \times 10^{-18}$). Additional terms highly enriched included epithelial cell proliferation (25 genes; adjusted $p = 1.75 \times 10^{-16}$), radiation response (24 genes; adjusted $p = 3.41 \times 10^{-16}$), and regulation of epithelial cell proliferation (24 genes; adjusted $p = 1.06 \times 10^{-16}$). Phosphatidylinositol 3-kinase signalling was significantly enriched (14 genes; adjusted $p = 1.09 \times 10^{-12}$), consistent with the inclusion of components of the PI3K pathway in the DRIVER_CORE set. Cellular senescence (12 genes; adjusted $p = 1.71 \times 10^{-11}$) and negative regulation of the cell cycle (20 genes; adjusted $p = 6.44 \times 10^{-13}$) were also among the terms most enriched, reflecting the representation of tumour suppressor mechanisms. The top 15 enriched GO terms are shown in Figure S1a, and the complete list is available in SM-GO.

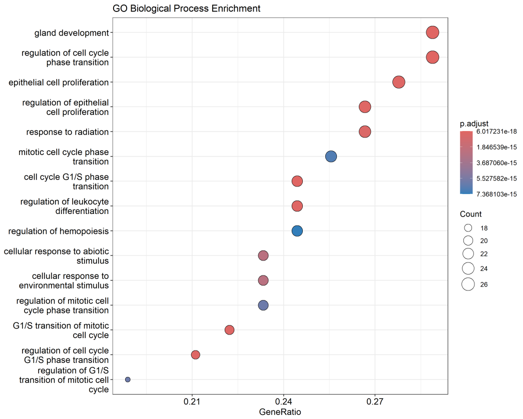


(a)

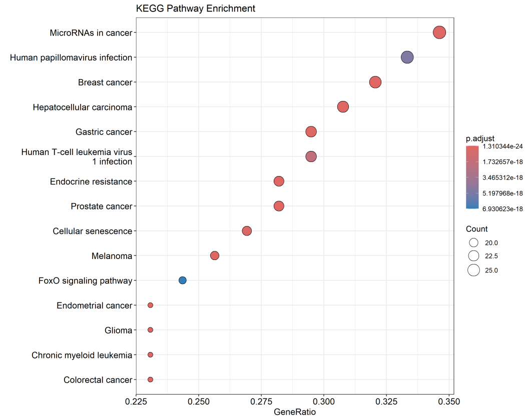


(b)

Figure S1: Pathway enrichment analysis of the DRIVER_CORE gene set. (a) Gene Ontology Biological Process enrichment. (b) KEGG pathway enrichment. Dot size indicates the number of genes; colour indicates adjusted p-value.

##### 1.2 KEGG Pathway Enrichment

KEGG pathway analysis identified 68 significantly enriched pathways (adjusted p $< 0.05$). The most significantly enriched pathway was Breast cancer (29 genes; adjusted p $= 1.31 \times 10^{-24}$), followed by Endocrine resistance (22 genes; adjusted p $= 1.79 \times 10^{-24}$) and the PI3K-Akt signalling pathway (27 genes; adjusted p $= 3.63 \times 10^{-16}$). Additional cancer-related pathways with significant enrichment included Proteoglycans in cancer (21 genes; adjusted p $= 1.12 \times 10^{-14}$), Prostate cancer (16 genes; adjusted p $= 6.93 \times 10^{-14}$), and Hepatocellular carcinoma (17 genes; adjusted p $= 3.63 \times 10^{-13}$). Enrichment was also observed for Cellular senescence (16 genes; adjusted p $= 5.96 \times 10^{-12}$), p53 signalling pathway (11 genes; adjusted p $= 3.03 \times 10^{-10}$), and Cell cycle (13 genes; adjusted p $= 1.06 \times 10^{-9}$). Cross-cancer enrichment was evident, with significant results for Melanoma (12 genes; adjusted p $= 1.02 \times 10^{-11}$), Gastric cancer (14 genes; adjusted p $= 1.71 \times 10^{-10}$), and Colorectal cancer (12 genes; adjusted p $= 3.85 \times 10^{-10}$). The top 15 enriched KEGG pathways are shown in Figure S1b, and the complete list is available in SM-KEGG.

### 2 Breast Cancer Prognosis

###### 2.0.1 In Distribution Experiment Results

Table S1: Benchmarking survival models across 12 datasets under the in-distribution (InD) setting.

| **Dataset** | **TabSurv** | **DeepHS** | **DeepSurv** | **LH** | **MTLR** | **PCHazard** | **PMF** | **RSF** |
|---|---|---|---|---|---|---|---|---|
| TCGA500 | **0.6251** | 0.4661 | 0.4097 | 0.4934 | 0.4701 | 0.5601 | 0.5538 | *0.5584* |
| GEO | 0.5578 | 0.5099 | 0.5474 | 0.5677 | 0.5784 | *0.6047* | 0.5660 | **0.6305** |
| GSE6532 | 0.6372 | *0.6549* | 0.6068 | 0.6553 | 0.5883 | 0.6513 | **0.6692** | 0.6551 |
| GSE19783 | 0.5556 | 0.5636 | **0.5790** | 0.5103 | 0.5275 | 0.4674 | *0.5670* | 0.5000 |
| HEL | **0.5775** | 0.5335 | 0.5866 | 0.5358 | 0.5196 | 0.4480 | 0.5081 | 0.5312 |
| unt | *0.6356* | 0.6022 | 0.5451 | 0.4586 | 0.4401 | 0.4843 | 0.5930 | **0.6593** |
| nki | **0.7295** | 0.5968 | 0.6425 | 0.6652 | 0.5971 | 0.6624 | 0.6243 | 0.6617 |
| transbig | 0.5912 | 0.5375 | 0.5871 | 0.6367 | 0.6001 | **0.6604** | 0.5918 | *0.6432* |
| UK | *0.6740* | 0.6335 | 0.6372 | 0.6593 | 0.6452 | **0.6941** | 0.6584 | 0.6198 |
| mainz | *0.7222* | **0.7841** | 0.6707 | 0.6390 | 0.5900 | 0.5867 | 0.6292 | 0.6848 |
| upp | 0.5396 | 0.5714 | 0.5828 | 0.5860 | *0.6353* | 0.6283 | **0.6416** | 0.6346 |
| METABRIC | **0.6218** | 0.5535 | *0.6125* | 0.5964 | 0.5945 | 0.5805 | 0.5732 | 0.5950 |

**Notes:** LH, Logistic Hazard; PMF, Probability Mass Function; DeepHS, DeepHitSingle; MTLR, Multi-Task Logistic Regression; RSF, Random Survival Forest. Best-performing methods are shown in bold; second-best methods are underlined.

For the in-distribution scenario, each dataset is partitioned using a single random split, with 60% of the samples allocated for training and 40% reserved for testing, using a fixed random seed of 42 to ensure reproducibility. Across the 12 datasets, Table S1 demonstrates that TabSurv achieves the highest C-index on 4 datasets and ranks second on 3 others, exhibiting competitive performance and consistent superiority over all baseline survival models.

Several baseline models, specifically LogisticHazard, MTLR, and PCHazard, exhibit noticeable instability, with C-index values falling below 0.5 on challenging datasets (e.g., *HEL*, *unt*). Such fluctuations are likely attributable to limited sample sizes and a reliance on restrictive parametric assumptions, which increase susceptibility to overfitting. In contrast, TabSurv maintains more consistent performance across the majority of datasets, potentially benefiting from the robust prior knowledge inherent in the foundation model and the proposed two-stage strategy for handling censored data.

### 3 Treatment Recommendation Evaluation

#### 3.1 Comparison of TabSurv Recommendation performance across 3 Genes sets

Table S2: Comparison of mean survival time (years) between the *Followed* and *Not Followed* groups across the three gene sets under the TabSurv recommendation framework.

| Genes Set | Mean survival (Followed Group) | Mean survival (Not Followed Group) | $\Delta$ Mean survival |
|---|---|---|---|
| Nik-94 genes | 16.32 | 12.45 | **3.86** |
| Mills-45 genes | 13.24 | 13.34 | −0.10 |
| Pereira-40 genes | 13.68 | 12.94 | 0.74 |

$\Delta$ Mean survival denotes the difference between *Followed* Recommendation and *Not Followed* Recommendation groups.

Table S2 summarises the mean survival time for patients who followed versus did not follow the TabSurv treatment recommendation across the three investigated gene sets. The *Nik-94 genes* set demonstrates the largest survival separation, with a mean survival of 16.32 years in the Followed group compared to 12.45 years in the Not Followed group, yielding a substantial difference of 3.86 years. In contrast, the *Pereira-40 genes* set shows only a modest survival improvement (0.74 years), while the *Mills-45 genes* set exhibits no meaningful separation between groups (-0.10 years). These results indicate that the *Nik-94 genes* DRIVER_CORE set provides the strongest discriminatory signal for treatment recommendation within the TabSurv framework.

#### 3.2 Kaplan-Meier Curve of TabSurv Recommendation and Baseline Methods Using All Gene

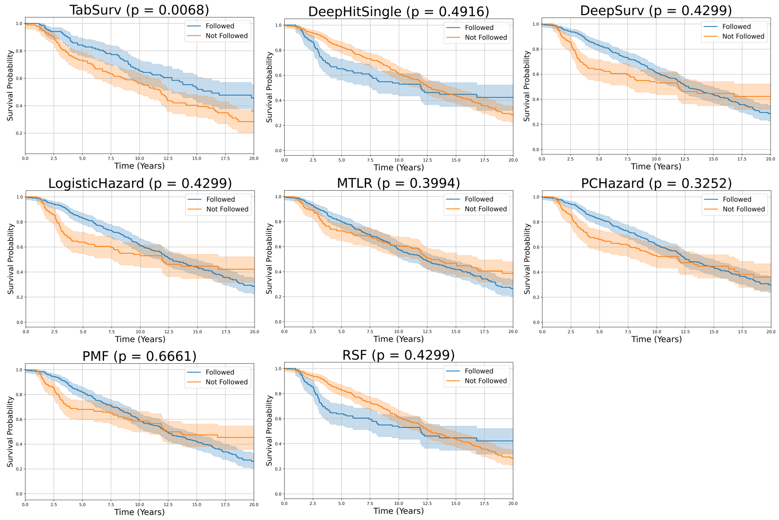


Figure S2: Kaplan–Meier survival curves comparing personalised treatment recommendations from TabSurv and baseline models using the full gene set. For each method, patients are stratified into *Followed* and *Not Followed* groups based on whether the received treatment matches the model-recommended therapy. Shaded areas represent 95% confidence intervals. Statistical significance between groups is assessed using the log-rank test, with corresponding $p$-values reported.

Figure S2 presents KM survival curves with 95% confidence intervals for seven survival models adapted for treatment recommendation. For each method, patients are stratified according to whether they followed the model's recommended treatment or not.

The log-rank $p$-value is reported as a measure of statistical separation between the Followed and Not Followed survival curves. While a small $p$-value indicates that the two survival curves are statistically distinct within a given time window, it does *not* by itself quantify the clinical benefit of a treatment recommendation. In particular, statistical significance does not guarantee that patients who followed the recommendation achieved longer survival; it only confirms that the survival distributions differ. Therefore, $p$-values should be interpreted as a supporting metric for curve separation rather than a direct measure of recommendation effectiveness.

#### 3.3 Kaplan-Meier Curve of TabSurv Recommendation with Baseline Methods Using Selected Genes

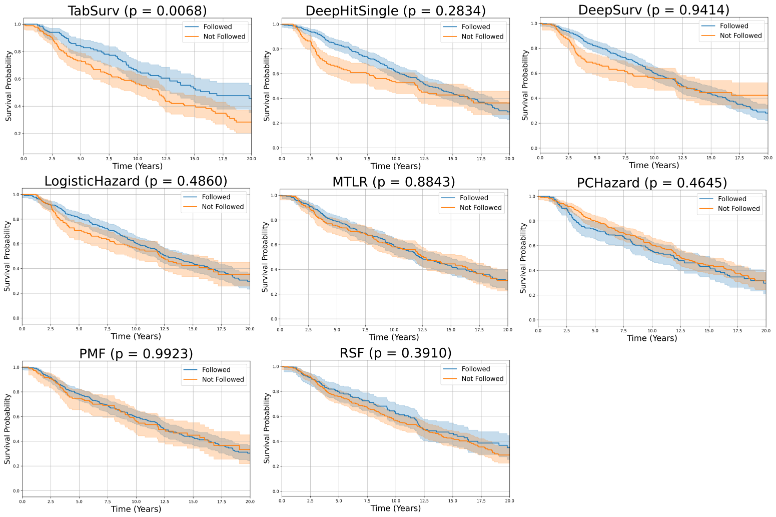


Figure S3: Kaplan–Meier survival curves for personalised treatment recommendation using selected cancer driver genes. For TabSurv and each baseline model, patients are stratified into *Followed* and *Not Followed* groups based on whether the treatment actually received matches the model-recommended therapy. Shaded areas represent 95% confidence intervals. Group differences are assessed using the log-rank test, with corresponding $p$-values reported.

Figure S3 presents Kaplan–Meier survival curves for all baseline methods when restricted to the curated set of biologically important genes, compared to the TabSurv method. The curves show that, for TabSurv, the Followed group consistently exhibits a higher survival probability than the Not Followed group, whereas the baseline methods do not demonstrate a clear long-term survival benefit up to the maximum follow-up time.

[3] Marc Carlson. *org.Hs.eg.db: Genome wide annotation for Human*, 2024. R package version 3.20.0.

[4] Minoru Kanehisa and Susumu Goto. Kegg: Kyoto encyclopedia of genes and genomes. *Nucleic Acids Research*, 28(1):27–30, 2000.
